# Prevalence and predictors of residual antibiotics in children’s blood in community settings in Tanzania

**DOI:** 10.1101/2023.08.30.23294851

**Authors:** Theopista Lotto, Sabine Renggli, Eliangiringa Kaale, Honorati Masanja, Beatrice Ternon, Laurent Arthur Décosterd, Valérie D’Acremont, Blaise Genton, Alexandra V. Kulinkina

## Abstract

**Introduction:** Children account for a significant proportion of antibiotic consumption in low- and middle-income countries, with the overuse of antibiotics occurring in both formal and informal health sectors. This study assessed the prevalence and predictors of residual antibiotics in children’s blood in Mbeya and Morogoro regions of Tanzania.

**Methods:** The cross-sectional community-based survey used two-stage cluster sampling to include 1,699 children under 15 years of age. For each child, information on recent illness, healthcare-seeking behavior, and the use of antibiotics, as well as a dried blood spot (DBS) sample, were collected. The samples underwent tandem mass spectrometry analysis to quantify the concentrations of 15 common antibiotics. Associations between survey variables and presence of residual antibiotics were assessed using mixed-effects logistic regression.

**Results:** The overall prevalence of residual antibiotics in the blood samples was 17.4% (95% CI: 15.6– 19.2), with the highest prevalence among under-five children. The most frequently detected antibiotics were trimethoprim (8.5%), sulfamethoxazole (6.0%), metronidazole (3.6%) and amoxicillin (2.5%). The strongest predictors of residual antibiotics in the blood were observed presence of antibiotics at home (aOR=2.9; 95% CI: 2.0–4.1) and reported consumption of antibiotics in the last two weeks (aOR=2.5; 95% CI: 1.6–3.9). However, half of the children who had residual antibiotics in their blood had no reported history of illness or taking antibiotics in the last two weeks, and antibiotics were not found in the home.

**Conclusion:** The study demonstrated high prevalence of antibiotic exposure among children in Tanzanian communities, albeit likely still underestimated, especially for compounds with short half-lives. A significant proportion of antibiotic exposure was unexplained and may be due to unreported self-medication or environmental pathways. Incorporating biomonitoring into surveillance strategies can help to better understand exposure patterns and design antibiotic stewardship interventions.

**Key messages:** *What is already known about this topic?:*

- Children are major consumers of antibiotics in low- and middle-income countries, worsening the emergence of antibiotic resistance.
- There is high reported use of prescribed and non-prescribed antibiotics in Tanzania, but no reliable data about the prevalence of antibiotic exposure in the pediatric population.

*What are the new findings?:*

- The prevalence of residual antibiotics in children’s blood is high, albeit still likely under-estimated, especially for compounds with short half-lives.
- Children are often exposed to several antibiotics concurrently, many with their exposure unexplained by illness history or reported consumption.

*Recommendations for policy:*

- Integrating objective quantification of antibiotic exposure into monitoring and surveillance strategies can help to better understand exposure patterns and design stewardship interventions.

## Introduction

Antibiotic overuse contributes to the rise in antimicrobial resistance, a serious problem for healthcare systems globally that threatens to reverse the gains made in extending life expectancy (1–4). In low- and middle-income countries (LMICs) that experience a high burden of infectious diseases, overconsumption of antibiotics occurs in both formal and informal health sectors, caused in part by a lack of regulation and by healthcare providers’ inability to distinguish between viral and bacterial infections (5–7). In the formal health sector, high use of antibiotics has been reported in inpatient (8) and outpatient (9) settings. Self-medication with antibiotics is also common, with a prevalence above 50% according to a systematic review of 40 African studies conducted between 2005 and 2020 (10). The main sources of non-prescribed antibiotics in LMICs are private pharmacies, informal medicine vendors, and medications stored at home (11). For example, in In Tanzania, up to 90% of drug shop owners report selling antibiotics based only on patients’ requests, regardless of symptoms (12,13).

Studies suggest that young children are major consumers of antibiotics in LMICs due to higher frequency of illnesses and health seeking behaviors (14,15). Despite the role of the Integrated Management of Childhood Illness protocol in improving antimicrobial prescribing practices in theory (16), in practice, health workers at the primary care level still lack sufficient clinical skills, pharmacological knowledge, and diagnostics (17). Consequently, over 60% of outpatient visits for children under the age of five years result in antibiotic prescriptions (18), often unnecessarily (9). Exposure to antibiotics in childhood is linked to a number of negative long-term health consequences, such as allergies, obesity, and neurodevelopment disorders (19).

While health facility-based antibiotic exposure surveys in LMICs are relatively common (20,21), community-based studies remain rare. Those that are available rely primarily on reported history of antibiotic consumption (14,22), resulting in a lack of reliable data about the prevalence of antibiotic exposure in the community. To address this gap, the aim of the present study was to establish a community-level prevalence of residual antibiotics in children’s blood and assess individual and household level risk factors of antibiotic exposure. Dried Blood Spot (DBS) samples were utilized to quantify residual antibiotics in the blood samples as an objective and rigorous way to monitor drug pressure in the population, compared to reported history used in prior studies.

## Methods

The study comprised of a cross-sectional household survey and DBS sample collection. Residual antibiotic concentrations were quantified in the samples, and results were analyzed together with individual and household-level predictors of exposure.

### Study setting

Tanzania has four administrative levels: regions, councils, wards, and villages (in rural areas) or streets (in urban areas). Villages are further divided into sub-villages, whereas streets are not. The survey was conducted in five councils of two regions: Mbeya City Council (CC) and Mbeya District Council (DC) in Mbeya, and Mlimba DC, Ulanga DC, and Ifakara Town Council (TC) in Morogoro (Figure 1). All study areas were rural, except Mbeya CC that was urban with a higher population density. Morogoro region generally experiences higher prevalence of malaria and acute respiratory infections and has slightly lower access to healthcare facilities as compared to Mbeya region (Table S1, Supplemental Information).

**Figure 1.**
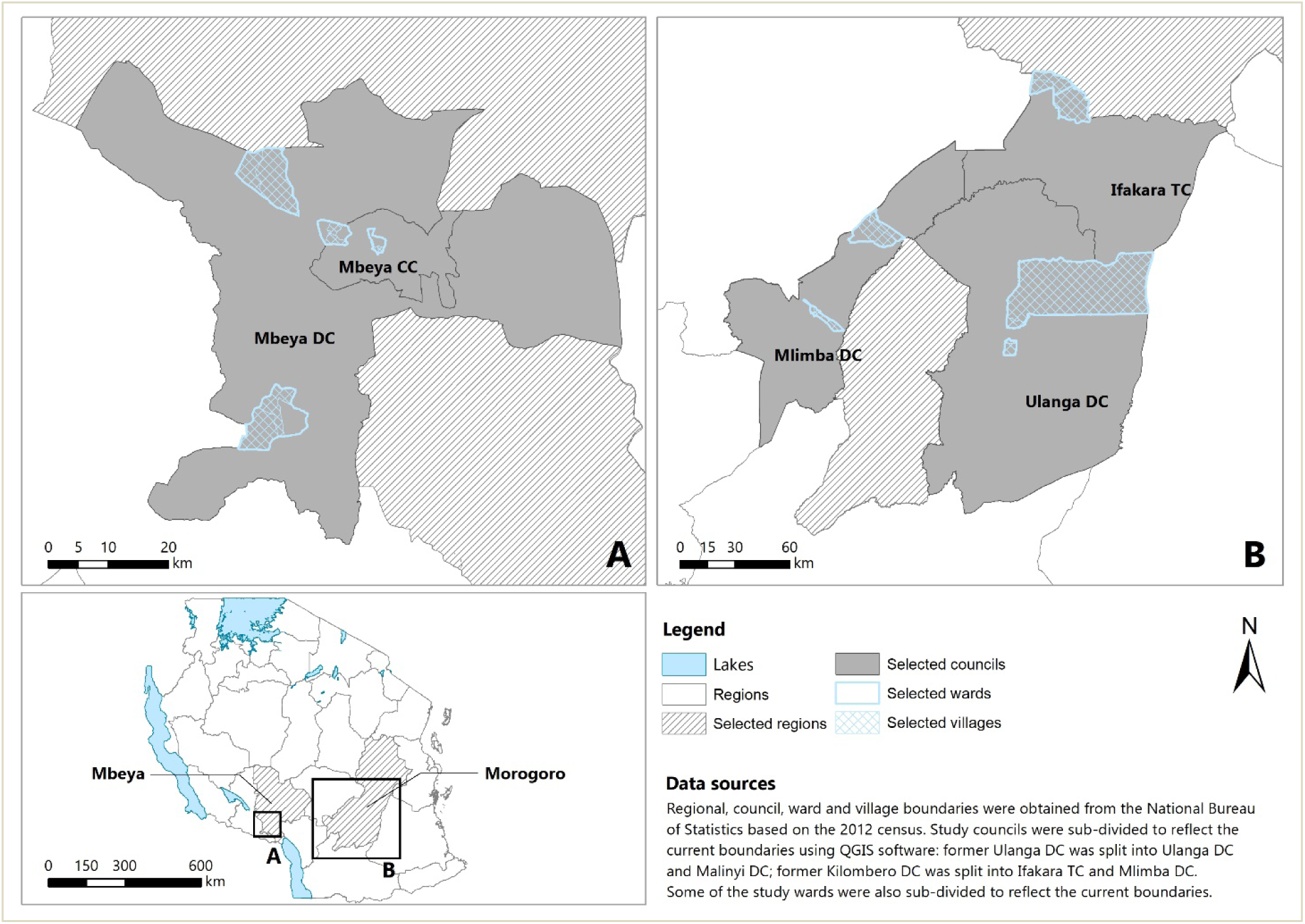
Map showing the locations of the ten wards (clusters) sampled as part of this community-based study. Data sources: administrative boundaries [[Tanzania National Bureau of Statistics](https://www.nbs.go.tz/index.php/en/census-surveys/gis)], lakes [[RCMRD geoportal](https://rcmrd.africageoportal.com/datasets/ba8eca5d9a5d47e79f39f0311881421e_0/explore?location=10.004711%2C-1.333777%2C9.00)].

### Study design and sample size

The cross-sectional survey employed a two-stage cluster sampling approach, with ward as the primary cluster and village (or street) as the secondary cluster. A total of 20 wards constituted the sampling frame: these were wards that contained a randomly selected primary health facility for a future intervention trial. Out of the 20 wards, 10 were purposively selected for the survey (Figure 1). Selection considered balance among study councils (two per council), malaria endemicity, and access to healthcare services. The expected prevalence of children with residual antibiotics in the blood was unknown. A feasible sample size of 1,700 children, approximately 170 children from each of the 10 wards, was sufficiently powered to measure a prevalence of 3% or higher (with a power of 80%, confidence level of 95%, and intracluster correlation coefficient of 0.05), calculated using the epiR package in R software (version 4.2.1).

### Sampling procedure

Selected wards were listed with their corresponding villages. Two villages were purposively selected from each ward based on proximity to the future intervention health facility. Within each village, half of the sub-villages were selected using a simple random sample. A total of 170 children were sampled per ward and allocated to villages proportionately to the number of children under 15 years of age estimated from the regional population profiles (23,24). The village sample was allocated to the sub-village level proportionately to the number of households provided by the ward executive officers (Table S2, Supplemental Information). Households in each sub-village were selected randomly. In each household, all children under 15 years of age whose caregiver consented to participate were included; those with chronic or severe conditions requiring regular hospital attendances were excluded. Although a true probability sample was not used, the sampled population represents average healthcare access in the study area (Figure S1, Supplemental Information).

### Field data collection

The survey was conducted between 1 February and 14 March 2021. The questionnaire was administered in Swahili on tablets using Open Data Kit (ODK). Household level data was collected first, followed by individual level data for each eligible child. Household variables included GPS location, socioeconomic characteristics, usual self-medication practices and healthcare seeking behaviors. For each child, basic demographics, acute illness in the 14 days prior to the survey, and healthcare seeking behavior for the corresponding illness were collected. A malaria rapid diagnostic test (mRDT) was performed on-site, and a DBS sample collected. Completed surveys were checked for accuracy and completeness daily and sent to the central server.

### Laboratory procedures

DBS samples were collected by finger prick, dried at room temperature for at least two hours, placed in a re-sealable plastic bag with desiccant, and stored at −10°C in the field for a maximum of 6 days. Samples were then transferred to the main research laboratories at National Institute for Medical Research – Mbeya Medical Research Center (NIMR–MMRC) and Ifakara Health Institute (IHI) and stored at −80°C before being shipped to Switzerland on dry ice for analysis. DBS samples were analyzed for 15 pre-selected antibiotics using liquid chromatography coupled with tandem mass spectrometry (LC-MS/MS), the method developed at the Lausanne University Hospital in Switzerland (25). The method was adapted to the needs of the survey and validated prior to use in this analysis. Quantified antibiotics included amoxicillin, ampicillin, cloxacillin, penicillin G, penicillin V, azithromycin, erythromycin, ciprofloxacin, ceftriaxone, cephalexin, chloramphenicol, doxycycline, sulfamethoxazole, trimethoprim, and metronidazole. Antibiotics were selected based on the Tanzania essential medicines list, local availability, and common use in the formal and informal health sectors (Table S3, Supplemental Information).

### Statistical analysis

Data was cleaned in Stata software (version 16.1) and transformed and analyzed in R software (version 4.1.2). Continuous variables were categorized and assessed for linearity with the response variable before inclusion in regression models. Differences in distributions of categorical variables were assessed using a χ^2^ test (26). Concordance between two binary proxy measures of antibiotic exposure and presence of antibiotics in the blood was assessed using a combination of a Chi-square (χ^2^) test and a Phi (ϕ) correlation coefficient.

A mixed-effects logistic regression model (glmer function in the lme4 package in R software) was used to identify factors associated with presence of antibiotics in the blood. Odds ratio (OR), 95% confidence interval (CI) and p-value were reported. Ward was included as a random effect in univariable and multivariable models to account for clustering of observations. Initially, all child- and household-level variables were considered for inclusion in the analysis, but some were excluded based on lack of relevance or limited sample size (Tables S4 and S5, Supplemental Information). Principal component analysis was used to reduce dimensionality of the socioeconomic variables (Table S6, Supplemental Information) and socioeconomic status (SES) quintile variable was analyzed. In total, 11 explanatory variables (Table 1) were included as fixed effects in the univariable analysis. Two variables were further excluded from the multivariable analysis due to high correlation with another included variable. All interactions were tested but excluded from the analysis to maintain interpretability of the results.

**Table 1.**
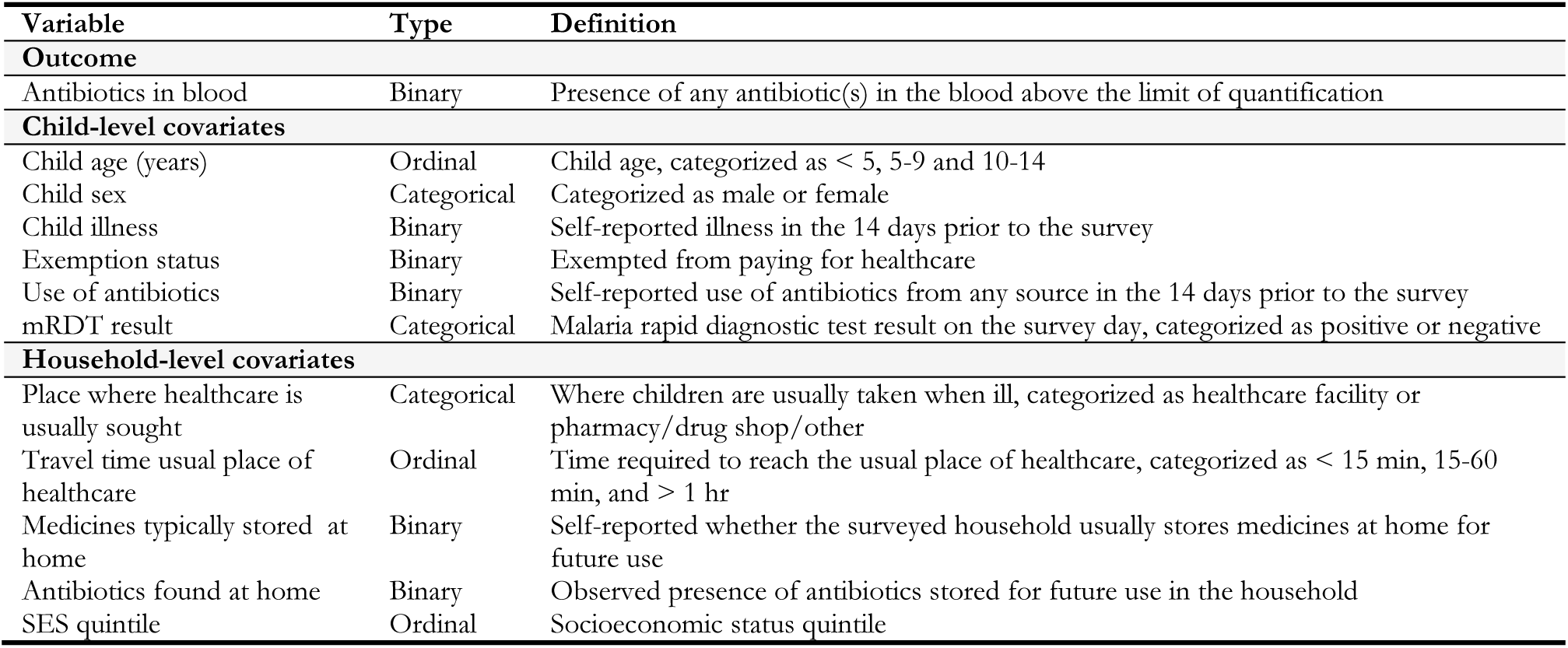
Summary of the primary analysis variables.

| Variable | Type | Definition |
| --- | --- | --- |
| **Outcome** |  |  |
| Antibiotics in blood | Binary | Presence of any antibiotic(s) in the blood above the limit of quantification |
| **Child-level covariates** |  |  |
| Child age (years) | Ordinal | Child age, categorized as < 5, 5-9 and 10-14 |
| Child sex | Categorical | Categorized as male or female |
| Child illness | Binary | Self-reported illness in the 14 days prior to the survey |
| Exemption status | Binary | Exempted from paying for healthcare |
| Use of antibiotics | Binary | Self-reported use of antibiotics from any source in the 14 days prior to the survey |
| mRDT result | Categorical | Malaria rapid diagnostic test result on the survey day, categorized as positive or negative |
| **Household-level covariates** |  |  |
| Place where healthcare is usually sought | Categorical | Where children are usually taken when ill, categorized as healthcare facility or pharmacy/drug shop/other |
| Travel time usual place of healthcare | Ordinal | Time required to reach the usual place of healthcare, categorized as < 15 min, 15-60 min, and > 1 hr |
| Medicines typically stored at home | Binary | Self-reported whether the surveyed household usually stores medicines at home for future use |
| Antibiotics found at home | Binary | Observed presence of antibiotics stored for future use in the household |
| SES quintile | Ordinal | Socioeconomic status quintile |

### Ethics statement

The ethical review boards of IHI (IHI/IRB/No: 20-2020) and NIMR (NIMR/HQ/R.8a/Vol. IX/3463) in Tanzania and the Ethikkommission Nordwest- und Zentralschweiz (AO_2020-00050) in Switzerland approved the original protocol and subsequent amendments. For each participating child, an adult household member (aged 18 years of above) provided written informed consent in Swahili. If they could not read or write, they were asked to find a witness, who signed in addition to confirm proper consent taking. Children above 11 years of age also provided written assent.

### Patient and public involvement

Patients and/or the public were not involved in the design, conduct, reporting or dissemination plans of this research.

## Results

During recruitment, 1668 households were approached and 1104 (66.2%) could have the eligibility criteria assessed. Reasons for not being able to asses household eligibility included inability to locate the household (n=309), no one being at home (n=167), and other reasons (n=88). Of the 1104 households, 294 (26.6%) did not have children under the age of 15 and were excluded. Of the remaining 810 households, 735 (91%) provided consent and were included in the study. The study households contained 1879 children; however, some were not present at the time of the survey (n=125) and another 12 declined to participate, resulting in 1742 children who were surveyed. Further, we excluded 43 children whose lab samples were mislabeled, misplaced, or of poor quality. The final dataset included 1699 children with complete survey data and a laboratory result (Figure 2).

**Figure 2.**
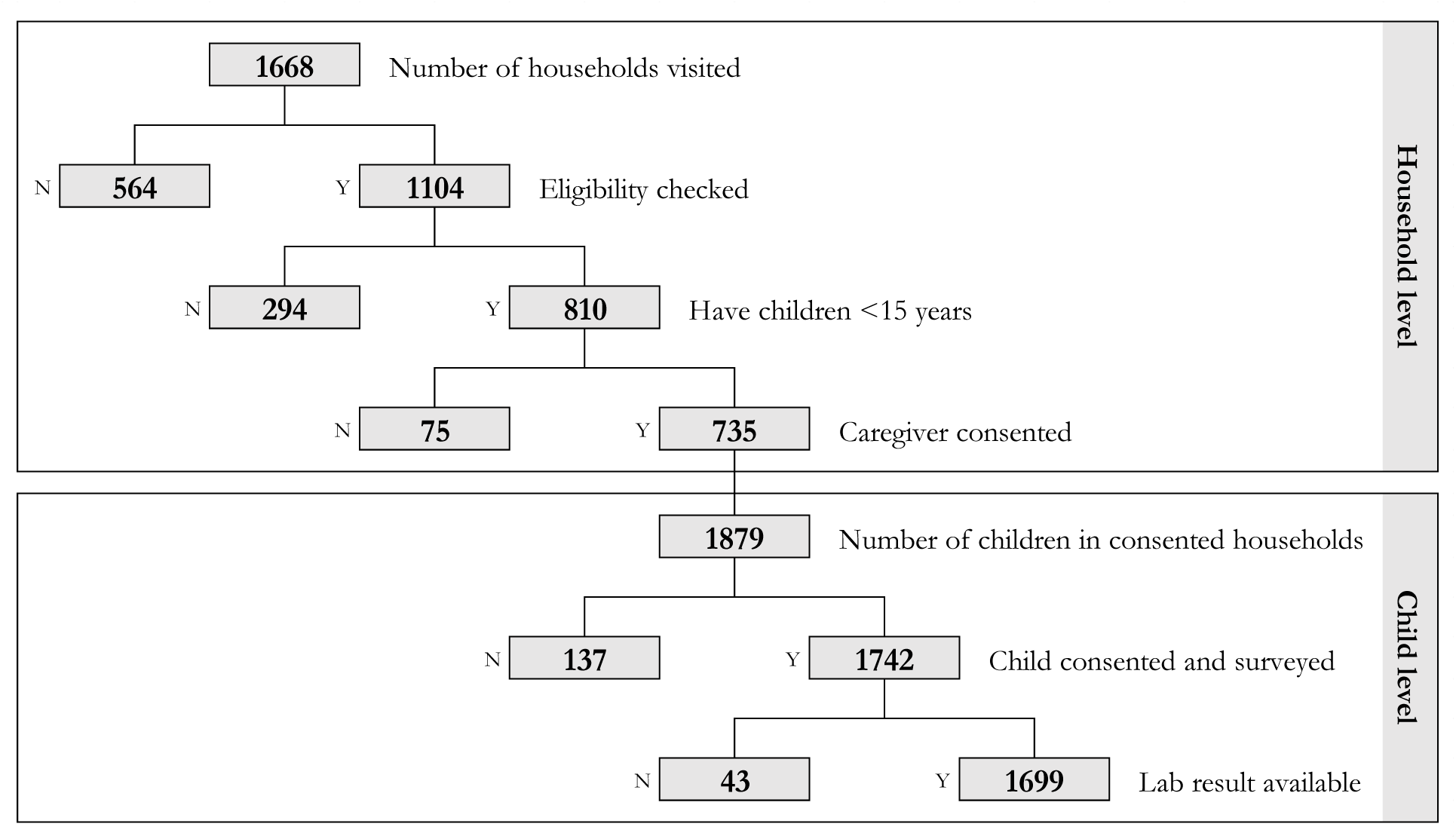
Flowchart showing the recruitment of study participants (N = No; Y = Yes).

Approximately 60% (1,010) of the sampled children were from Morogoro and 40% (689) from Mbeya regions (Table 2). There was an equal sex distribution, and the predominant age group of children was under 5 years (43%). Over 90% of participants lived in households where a health facility was the usual place to seek care in case of illness. However, households in Morogoro reported longer travel times to their usual place of healthcare and generally had lower SES (Table 2). Approximately 30% of children had been sick within 14 days of the survey, and 9% reported having taken an antibiotic. Of all surveyed children, 27% in Morogoro and none in Mbeya had a positive mRDT result. However, Mbeya had a higher overall prevalence of illness history (34.1% versus 27.1%) and reported consumption of antibiotics (11.3% versus 7.2%) as compared to Morogoro (p=0.004). Self-reported storage of medicines (31.3% versus 18.2%) and observed presence of antibiotics at home (15.5% versus 8.8%) were also higher in Mbeya (p<0.001). A total of 296 children had at least one antibiotic detected in their blood, with an overall prevalence of 17.4% (95% CI: 15.6–19.2), with no significant difference between Mbeya (18.6%) and Morogoro (16.7%) regions (Table 2).

**Table 2.**
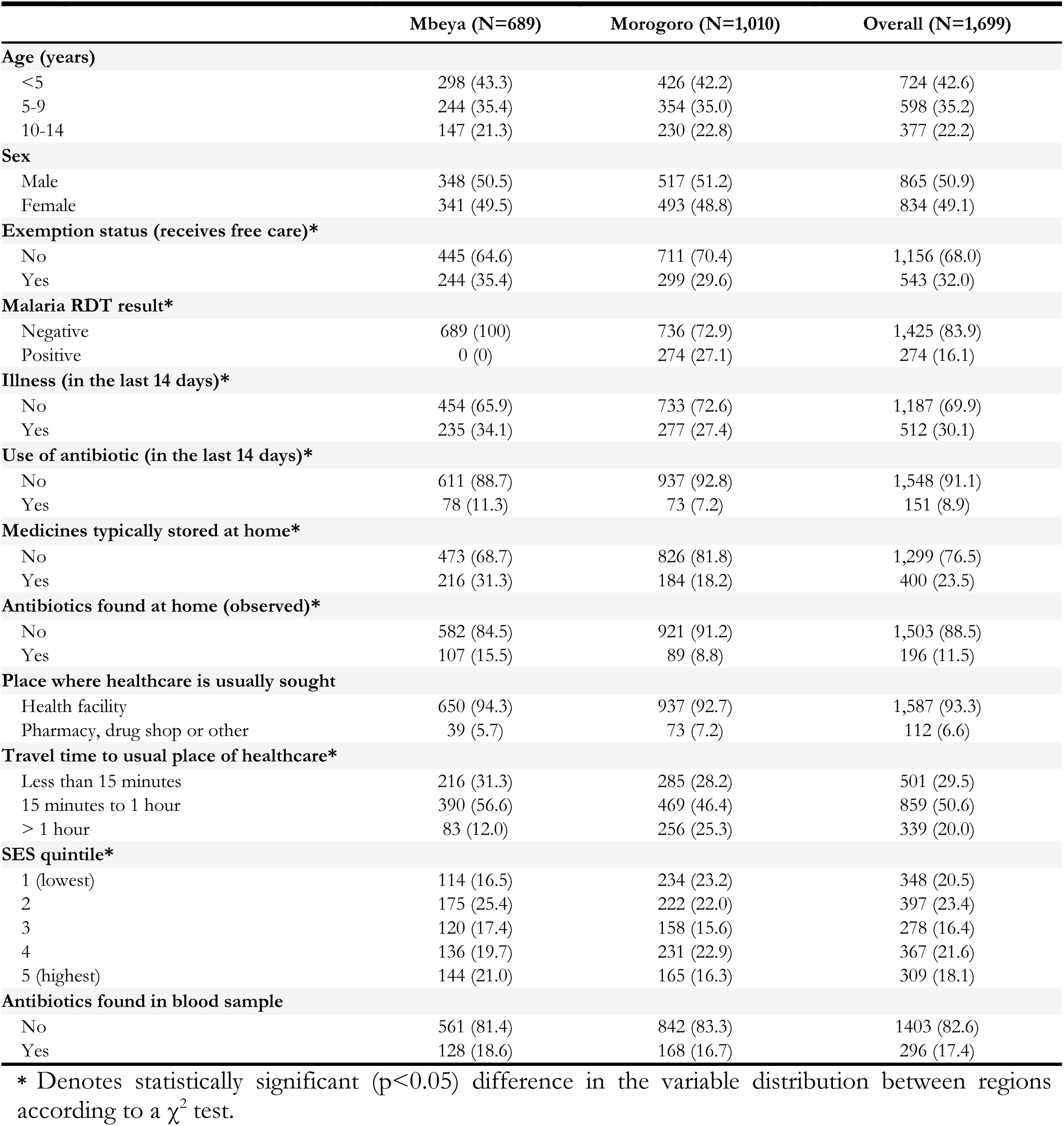
Characteristics – n (%) of study participants (children below 15 years of age).

All 15 antibiotics were detected at least once, with the most prevalent being trimethoprim (8.5%) and sulfamethoxazole (6%), representing exposure to co-trimoxazole (Table 3). Lab-quantified antibiotic presence in the blood was compared with two proxy measures of exposure assessed during the survey: observed storage of antibiotics in the home and caregiver-reported consumption of antibiotics by the child. Overall, there were weak but statistically significant correlations between presence of an antibiotic in the blood and observed storage of antibiotics at home (ϕ = 0.19, p<0.001) and reported consumption of an antibiotic (ϕ = 0.17, p<0.001) (Table 3). Correlation values generally tended to be higher for antibiotics with longer half-lives (e.g., trimethoprim, sulfamethoxazole, metronidazole, azithromycin) as compared to those with short half-lives (e.g., amoxicillin, ciprofloxacin, cloxacillin) (Table 3).

**Table 3.**
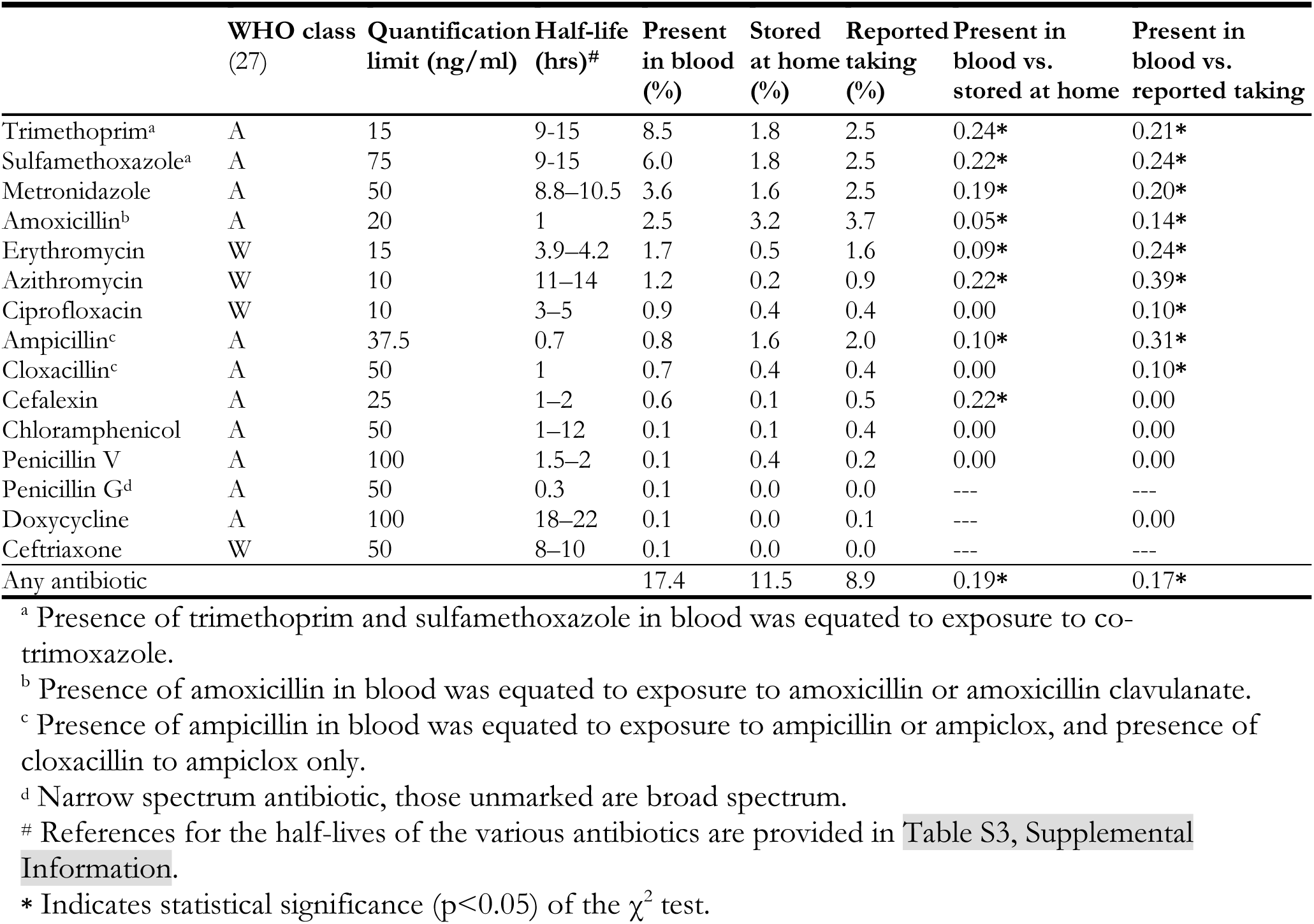
Prevalence of 15 antibiotics quantified in blood samples, found stored at home during the survey, and consumed by the child in the last 14 days according to the caregiver. Phi correlation coefficients (ϕ) between presence in blood and the two survey variables are also presented.

|  | WHO class<br>(27) | Quantification<br>limit (ng/ml) | Half-life<br>(hrs)<sup>#</sup> | Present<br>in blood<br>(%) | Stored<br>at home<br>(%) | Reported<br>taking<br>(%) | Present in<br>blood vs.<br>stored at home | Present in<br>blood vs.<br>reported taking |
| --- | --- | --- | --- | --- | --- | --- | --- | --- |
| Trimethoprim<sup>a</sup> | A | 15 | 9-15 | 8.5 | 1.8 | 2.5 | 0.24* | 0.21* |
| Sulfamethoxazole<sup>a</sup> | A | 75 | 9-15 | 6.0 | 1.8 | 2.5 | 0.22* | 0.24* |
| Metronidazole | A | 50 | 8.8-10.5 | 3.6 | 1.6 | 2.5 | 0.19* | 0.20* |
| Amoxicillin<sup>b</sup> | A | 20 | 1 | 2.5 | 3.2 | 3.7 | 0.05* | 0.14* |
| Erythromycin | W | 15 | 3.9-4.2 | 1.7 | 0.5 | 1.6 | 0.09* | 0.24* |
| Azithromycin | W | 10 | 11-14 | 1.2 | 0.2 | 0.9 | 0.22* | 0.39* |
| Ciprofloxacin | W | 10 | 3-5 | 0.9 | 0.4 | 0.4 | 0.00 | 0.10* |
| Ampicillin<sup>c</sup> | A | 37.5 | 0.7 | 0.8 | 1.6 | 2.0 | 0.10* | 0.31* |
| Cloxacillin<sup>c</sup> | A | 50 | 1 | 0.7 | 0.4 | 0.4 | 0.00 | 0.10* |
| Cefalexin | A | 25 | 1-2 | 0.6 | 0.1 | 0.5 | 0.22* | 0.00 |
| Chloramphenicol | A | 50 | 1-12 | 0.1 | 0.1 | 0.4 | 0.00 | 0.00 |
| Penicillin V | A | 100 | 1.5-2 | 0.1 | 0.4 | 0.2 | 0.00 | 0.00 |
| Penicillin G<sup>d</sup> | A | 50 | 0.3 | 0.1 | 0.0 | 0.0 | --- | --- |
| Doxycycline | A | 100 | 18-22 | 0.1 | 0.0 | 0.1 | --- | 0.00 |
| Ceftriaxone | W | 50 | 8-10 | 0.1 | 0.0 | 0.0 | --- | --- |
| Any antibiotic |  |  |  | 17.4 | 11.5 | 8.9 | 0.19* | 0.17* |

<sup>a</sup> Presence of trimethoprim and sulfamethoxazole in blood was equated to exposure to cotrimoxazole.
<sup>b</sup> Presence of amoxicillin in blood was equated to exposure to amoxicillin or amoxicillin clavulanate.
<sup>c</sup> Presence of ampicillin in blood was equated to exposure to ampicillin or ampiclox, and presence of cloxacillin to ampiclox only.
<sup>d</sup> Narrow spectrum antibiotic, those unmarked are broad spectrum.
<sup>#</sup> References for the half-lives of the various antibiotics are provided in Table S3, Supplemental Information.
\* Indicates statistical significance ( $p < 0.05$ ) of the $\chi^2$ test.

Under-five children had the highest prevalence (22%) of antibiotics in their blood. Ages 5-9 and 10-14 years (Table 4) were associated with nearly 40% lower odds of antibiotic exposure as compared to the under-5 reference group in both unadjusted and adjusted regression models (Table 4). Self-reported consumption of antibiotics in the last 14 days (aOR = 2.47) and observed storage of antibiotics at home (aOR = 2.86) increased the odds of having antibiotics in the blood as compared to the respective reference groups (p<0.001). For both variables, unadjusted ORs were higher (3.36 and 3.32, respectively). Travel time to the usual place of healthcare (normally a health facility) of more than 1 hour was associated with 35% lower odds of having antibiotics in the blood as compared to the reference category of less than 15 minutes in adjusted (p=0.044) and unadjusted (p=0.034) models. The multivariable model explained 12.8% of the variability in the outcome, of which the fixed effects explained 9.6%, and the cluster random effect explained the additional 3.2%.

**Table 4.** Results of the univariable and multivariable mixed-effects logistic regression models explaining the presence of residual antibiotic in the blood samples of the study participants (N=1,699).

|  | Antibiotic prevalence<br>n / N (%) | Univariable<br>OR (95% CI)* | p-value | Multivariable<br>aOR (95% CI)* | p-value |
| --- | --- | --- | --- | --- | --- |
| **Age (years)** |  |  |  |  |  |
| <5 | 159 / 724 (22.0) | 1.00 |  | 1.00 |  |
| 5-9 | 85 / 598 (14.2) | 0.59 (0.44–0.80) | **<0.001** | 0.65 (0.42–0.87) | **0.006** |
| 10-14 | 44 / 377 (13.8) | 0.57 (0.40–0.80) | **0.001** | 0.61 (0.48–0.88) | **0.007** |
| **Sex** |  |  |  |  |  |
| Male | 147 / 865 (17.0) | 1.00 |  | 1.00 |  |
| Female | 149 / 834 (17.9) | 1.05 (0.82–1.35) | 0.700 | 1.12 (0.86–1.47) | 0.383 |
| **Exemption status (receives free care)** |  |  |  |  |  |
| No | 175 / 1156 (15.1) | 1.00 |  | --- | --- |
| Yes | 121 / 543 (22.3) | 1.65 (1.27–2.15) | **<0.001** |  |  |
| **Malaria RDT result** |  |  |  |  |  |
| Negative | 255 / 1425 (17.9) | 1.00 |  | 1.00 |  |
| Positive | 41 / 274 (15.0) | 0.98 (0.62–1.55) | 0.946 | 1.47 (0.91–2.38) | 0.114 |
| **Illness (in the last 14 days)** |  |  |  |  |  |
| No | 182 / 1187 (15.3) | 1.00 |  | 1.00 |  |
| Yes | 114 / 512 (22.3) | 1.59 (1.22–2.07) | **0.001** | 0.93 (0.66–1.30) | 0.663 |
| **Consumption of antibiotic (in the last 14 days)** |  |  |  |  |  |
| No | 238 / 1548 (15.4) | 1.00 |  | 1.00 |  |
| Yes | 58 / 151 (38.4) | 3.36 (2.33–4.83) | **<0.001** | 2.47 (1.57–3.88) | **<0.001** |
| **Medicines typically stored at home (reported)** |  |  |  |  |  |
| No | 193 / 1299 (14.9) | 1.00 |  | --- | --- |
| Yes | 903 / 400 (25.8) | 1.89 (1.43–2.50) | **<0.001** |  |  |
| **Antibiotics found at home (observed)** |  |  |  |  |  |
| No | 223 / 1503 (14.8) | 1.00 |  | 1.00 |  |
| Yes | 73 / 196 (37.2) | 3.32 (2.38–4.63) | **<0.001** | 2.86 (1.99–4.09) | **<0.001** |
| **Place where healthcare is usually sought** |  |  |  |  |  |
| Health facility | 284 / 1587 (17.9) | 1.00 |  | 1.00 |  |
| Pharmacy, drug shop or other | 12 / 112 (10.7) | 0.61 (0.33–1.13) | 0.115 | 0.53 (0.27–1.01) | 0.053 |
| **Travel time to usual place of healthcare** |  |  |  |  |  |
| Less than 15 minutes | 99 / 501 (19.8) | 1.00 |  | 1.00 |  |
| 15 minutes to 1 hour | 150 / 859 (17.5) | 0.85 (0.64–1.13) | 0.260 | 0.84 (0.62–1.13) | 0.248 |
| > 1 hour | 47 / 339 (13.9) | 0.65 (0.44–0.97) | **0.034** | 0.65 (0.43–0.99) | **0.044** |
| **SES quintile** |  |  |  |  |  |
| 1 (lowest) | 46 / 348 (13.2) | 1.00 |  | 1.00 |  |
| 2 | 60 / 397 (15.1) | 1.07 (0.69–1.65) | 0.767 | 0.95 (0.60–1.50) | 0.834 |
| 3 | 55 / 278 (19.8) | 1.52 (0.97–2.39) | 0.070 | 1.23 (0.76–2.00) | 0.397 |
| 4 | 73 / 367 (19.9) | 1.53 (0.99–2.37) | **0.057** | 1.46 (0.92–2.33) | 0.108 |
| 5 (highest) | 62 / 309 (20.1) | 1.74 (1.08–2.83) | **0.024** | 1.46 (0.88–2.43) | 0.147 |

\* Odds ratio (OR) or adjusted odds ratio (aOR) and 95% confidence interval (CI) for fixed effects are shown. All models also included cluster as a random effect.

History of illness and high SES (4^th^ and 5^th^ quintile) increased the odds of having antibiotics in the blood in the univariable models but lost significance in the multivariable model. On the other hand, lower odds of antibiotic exposure resulting from usually seeking care in a pharmacy or drug shop (as compared to a health facility) became nearly significant (p=0.053) in the adjusted model. Exemption status and typical practice of storing medicines at home were positively associated with antibiotic exposure in the univariable models but were excluded from the multivariable model due to being highly correlated with age and observed presence of antibiotics in the home, respectively. Sex and mRDT result were not significant in the univariable models but were retained in the multivariable model for potential comparability with other studies that control for these variables.

Of the 296 children who had antibiotics in their blood, 171 (58%) had one antibiotic, 98 (33%) two, and 27 (9%) three or more. The maximum number of antibiotics detected in a child’s blood sample was seven. Of the 27 children with three or more antibiotics in their blood, 24 had trimethoprim and sulfamethoxazole. Co-occurrence of trimethoprim, sulfamethoxazole and metronidazole was common (9/27). Only about half of these children had been ill (15/27) in the last 14 days and sought care (12/27), primarily in health facilities. Seven (25%) children, including the child with seven antibiotics in the blood sample, were not ill, did not report taking antibiotics, and did not have antibiotics stored in their home (Table S7, Supplemental Information). In fact, in the overall study population nearly half (145/296, 49%) of the children with antibiotics in their blood had this profile – i.e., no significant risk factors.

Lastly, predictors of antibiotic exposure among the subsets of children who had experienced illness in the 14 days prior to the survey (n=512) and subsequently those who sought care for the illness (n=312) were also assessed (Table S8, Supplemental Information). Of the 512 children who were ill, the majority (40%) reported respiratory symptoms only, with a higher percentage in Mbeya (52%) as compared to Morogoro (30%) (p<0.001). Approximately 61% of children who reported illness sought care, with no difference between regions. Of the 312 children who sought care, approximately 63% did so in health facilities, and the rest in drug shops and pharmacies, with no difference between regions. Malaria positivity, as determined while seeking care, was higher in Morogoro region (41/74) as compared to Mbeya (2/17). The rate of antibiotic prescription while seeking care was lower in Morogoro (36%) than in Mbeya (51%) (p=0.008). The most common antibiotics that resulted from seeking care were amoxicillin, co-trimoxazole, and metronidazole (Table S4, Supplemental Information). According to the results of mixed effects logistic regression models, among children who sought care, those who reported to have received antibiotics during care-seeking had approximately twice (aOR=2.02; 95% CI: 1.15–3.54) the odds of having antibiotics in their blood as compared to those who did not receive antibiotics. All other variables associated with illness and care-seeking were not significant (Table S8, Supplemental Information).

## Discussion

This study presents community-level prevalence and predictors of residual antibiotics in the blood of children under 15 years of age in Mbeya and Morogoro regions of Tanzania. To the best of our knowledge, this is the first community-based survey that quantified drug concentrations of a large number of antibiotics using a highly sensitive laboratory method. All 15 antibiotics tested were detected in DBS samples, with an overall prevalence of 17.4%. The most common antibiotics were trimethoprim, sulfamethoxazole, metronidazole, and amoxicillin. These trends are in line with recent data from the Tanzania Medicines and Medical Devices Authority reporting doxycycline, amoxicillin and trimethoprim-sulfamethoxazole as the three most frequently consumed antibiotics in 2017–2019, accounting for 20.01, 16.8 and 12.4 defined daily doses per 1000 inhabitants per day, respectively (28). These numbers are especially concerning since high rates of resistance to co-trimoxazole have been reported in sub-Saharan Africa and in Tanzania specifically (29,30). In this study, doxycycline was very rarely detected, probably because it is not recommended for children under the age of eight years. However, several broad-spectrum antibiotics (e.g., erythromycin, azithromycin, ciprofloxacin and ceftriaxone) belonging to the ‘Watch’ group of the WHO AWaRe Classification were detected. These drugs have higher resistance potential, and are thereby prioritized as key targets for monitoring and stewardship programs (27).

The main risk factors of antibiotic exposure were younger age, self-reported antibiotic consumption, observed antibiotic storage in the home, and shorter travel time to the usual place of receiving healthcare services. Prior Tanzanian studies have reported storage of antibiotics at home as a common practice (31,32). In our study, the most commonly stored antibiotics were amoxicillin, metronidazole and co-trimoxazole, similar to the findings of others (31,33). Our findings of generally better access to healthcare facilities and higher SES (albeit only in univariable analysis) being associated with higher antibiotic exposure agree with those of a Brazilian study (34). However, a negative mRDT test result did not increase antibiotic exposure, contrary to the findings of a recent meta-analysis (35). In fact, in our adjusted analysis, it was rather a positive mRDT result that was nearly significantly associated with increased odds of having antibiotics in the blood, probably because of easier general access to medicines. The practice of prescribing antibiotics to children with malaria in the study area while mRDTs are widely used warrants further investigation.

Some children had several (up to 7) antibiotics in their blood (Table S7, Supplemental Information). This may lead to adverse drug reactions (36), unfavorable health consequences later in life (19), and potentiation of microorganisms’ resistance to common antibiotics due to increased drug selection pressure (37). Another study in China also found up to six antibiotics in the urine samples of healthy primary school children (38). Notably, in our study, 49% of children who had antibiotics in their blood had not been ill, did not report taking antibiotics and no antibiotics were found in their home. Further investigation of these findings is needed to determine if there is substantial under-reporting of risk factors or potential significant exposure from environmental sources. Since several of the detected antibiotics are also used in the agricultural sector in Africa and in Tanzania specifically (39–41), and agriculture is a main economic activity in the study area, the possibility of environmental sources of exposure cannot be excluded.

In the present study, the overall correlation between self-reported consumption of antibiotics and antibiotic presence in the blood was rather weak (0.19). Correlation values with self-reporting tended to be higher for antibiotics with longer half-lives as compared to those with shorter half-lives, which is expected, reflecting underestimates of the prevalence of antibiotics with short half-lives that may have been eliminated by the time the blood samples were collected. Two other studies reported high correlation (0.72) between self-reported consumption and presence of antibiotics in the urine of patients with suspected enteric fever presenting to tertiary hospitals (21) and 85% concordance between same-day mother-reported antibiotic use and antibiotic prescription as documented on the medical care report form (14). However, these studies are not comparable to our study due to inclusion of a relatively healthy population of children in the community, up to 14-day recall period, and a different antibiotic quantification method. A study using similar methods investigate the relationship between self-reporting and presence of antimlarials in DBS samples found similarly poor correlation (42).

The present study has several strengths and potential limitations. To our knowledge, it is the first community-based study with a high number of antibiotics objectively measured in blood samples. While we measured 15 of Tanzania’s most commonly used antibiotics, a few others may have been missed. For example, three study participants reported taking gentamicin, but it was not included in the lab analysis. In the extensive survey of risk factors, a relatively long recall period of 14 days was used, which may have led to a higher correlation between reported risk factors and antibiotics with relatively long half-lives and lower correlation with those with short half-lives. In fact, the overall prevalence of antibiotics could be underestimated because most of the quantified antibiotics have relatively short half-lives as compared to the recall period. In future studies, it would be useful to ask about the timing of self-reported risk factors to further explore the relationship between under-reporting and under-detection of antibiotics with short half-lives and calibrate the overall prevalence through modeling. Lastly, the survey took place during the COVID-19 pandemic, with potentially atypical distribution of symptoms and antibiotics. However, we don’t expect that the pediatric population in our sample was significantly affected. Overall, we believe that the results of the study are generalizable to similar settings.

## Conclusion

Our study demonstrated high prevalence of antibiotic exposure in the pediatric population in the community, albeit likely still underestimated. Many children had several antibiotics in their blood, including broad-spectrum antibiotics in the ‘Watch’ group of the WHO AWaRe classification, which risks development of antimicrobial resistance due to increased drug selection pressure. A significant proportion of antibiotic exposure in our study is unexplained and may be due to unreported self-medication or environmental pathways. We recommend the use of objective measurements of antibiotic exposure in DBS samples in further studies and monitoring approaches, especially for antibiotics with longer half-lives. On the other hand, developing more precise self-reporting instruments for antibiotics with short half-lives may be warranted. Together, these complementary approaches can yield better estimates of overall antibiotic exposure. Lastly, we recommend considering the one health approach in questionnaires to integrate animal, environmental, and human perspectives. This is important for identifying the various pathways by which exposure occurs and for broadening the efforts to mitigate antibiotic resistance.

## Data Availability

All data produced in the present study are available upon reasonable request to the authors

## Acknowledgments

The authors wish to thank all research participants and government entities that facilitated the researchers’ access to the study sites. The authors also acknowledge the support of field staff in data collection, laboratory staff at the Lausanne University Hospital in analyzing the samples, as well as Prof. Maria Katapodi and students in her writing class for reviewing early versions of the manuscript. The authors would like to acknowledge the contribution of late Dr. Irene Masanja to the development of the study protocol prior to her passing in May 2020. Lastly, we acknowledge the financial support of the Swiss National Science Foundation (grant #179273).

## Contributors

BG and VDA conceptualized the project and acquired the funding. TL, SR, and AVK developed the study protocol and data collection plan. TL and SR collected the data and performed data quality assurance. TL, BT and LAD conducted the laboratory analyses. TL and AK conducted the statistical analysis and drafted the manuscript. BG, VDA, HM and EK provided critical review of the manuscript.

## Patient consent

The parent or primary caregiver of all study participants signed a consent form prior to being recruited into the study.

## Ethics approval

Ifakara Health Institute (IHI/IRB/No: 20-2020), National Institute for Medical Research (NIMR/HQ/R.8a/Vol. IX/3463) and the Ethikkommission Nordwest- und Zentralschweiz (AO_2020-00050).

## Data sharing statement

Requests for data access should be directed to the corresponding author and will be granted subject to approval by the National Institute for Medical Research of Tanzania.

