## Supplemental Material for "Prevalence and predictors of residual antibiotics in children’s blood in community settings in Tanzania"

#### Supplemental Information

**Table S1.** Characteristics of the study councils

**Table S2.** Sampled villages with corresponding population of children under 15 years of age

**Table S3.** Characteristics of antibiotics available in Tanzania

**Table S4.** Child-level variables that were measured in the survey

**Table S5.** Household-level variables that were measured in the survey (excluding SES variables)

**Table S6:** Household-level variables that were considered in the SES analysis

**Table S7.** Summary of 27 children who had three or more antibiotics in their blood

**Table S8.** Univariable and multivariable regression model results for children who reported illness

**Figure S1.** Map of the study area comparing access to health facilities among wards

**References**

**Table S1.** Characteristics of the study councils.

| Characteristics | Ref | Mbeya CC | Mbeya DC | Mlimba DC | Ifakara TC | Ulanga DC |
|---|---|---|---|---|---|---|
| Population[a] | (1) | 537,521 | 359,012 | 224,888 | 281,336 | 201,399 |
| Area (sq. km)[b] | (1) | 252 | 2,813 | 6,339 | 7,205 | 15,751 |
| Population density (per sq. km)[c] | -- | 2,133 | 128 | 35 | 39 | 13 |
| Number of administrative wards | (2, 3) | 36 | 28 | 16 | 19 | 21 |
| Number of villages (or streets) | (2, 3) | 181 | 152 | 62 | 81 | 59 |
| Problems in accessing health care due to distance (%)[d] | (4) | 42.4 | 42.4 | 51.6 | 51.6 | 51.6 |
| Number of operating health facilities[e] | (5) | 7/5/43 | 3/5/74 | 0/3/40 | 5/8/25 | 1/2/28 |
| Average household size | (6) | 4.2 | 4 | 4.3 | 4.3 | 4.9 |
| Malaria prevalence in under five children according to the rapid test (%)[d] | (7) | 4 | 4 | 9.5 | 9.5 | 9.5 |
| Acute respiratory infections (%)[d] | (4, 8) | 2.6 | 2.6 | 4.8 | 4.8 | 4.8 |
| Self-reported prevalence of diarrhea in the last 2 weeks among under five years (%)[d] | (4) | 13.9 | 13.9 | 8.5 | 8.5 | 8.5 |
| Average annual rainfall (mm) [d] | (9) | 1731.9 | 1731.9 | 1138.1 | 1138.1 | 1138.1 |

[a] Population was calculated by extrapolating village-level population of 2012 with the council level growth rate, per annum for the 2002-2012 intra-censal period, up to year 2021. Former Ulanga DC (2012) was split into Ulanga DC and Malinyi DC (not included here). Former Kilombero DC (2012) were split into Ifakara TC and Mlimba DC.

[b] Obtained from 2012 census shapefiles, subdivided further to reflect current council boundaries as explained above, using QGIS.

[c] Calculated by dividing population by area.

[d] Regional figure.

[d] Hospitals / health centers / dispensaries.

**Table S2.** Sampled villages with corresponding population of children under 15 years of age (as of 2020) based on data obtained from district planning officers.

| Village/street | # of sub-villages | 2020 pop[a] (1) | % pop children <15[a] (1) | 2020 pop <15 | Sampled children | Sampled sub-villages | Nearby health facilities (names) |
|---|---|---|---|---|---|---|---|
| **Morogoro region** | | | | | | | |
| **Ulanga DC** | | | | | | | |
| Ebuyu | 3 | 3065 | 43.6 | 1336 | 105 | 2 | 1 (Ebuyu) |
| Euga | 4 | 1901 | 43.6 | 829 | 66 | 2 | 1 (Ebuyu) |
| Lukande | 4 | 2003 | 43.6 | 873 | 87 | 2 | 1 (Lukande) |
| Gombe | 4 | 1954 | 43.6 | 852 | 84 | 2 | 1 (Lukande) |
| **Ifakara TC** | | | | | | | |
| Kidatu B | 3 | 5606 | 43.6 | 2444 | 88 | 2 | 2 (Kidatu & Tanesco) |
| Kidatu Kati | 3 | 5295 | 43.6 | 2309 | 83 | 2 | 2 (Kidatu & Tanesco) |
| Msolwa Stesheni | 2 | 5239 | 43.6 | 2284 | 87 | 1 | 2 (Msolwa Station & Msolwa game) |
| Nyange | 4 | 5034 | 43.6 | 2195 | 84 | 2 | 2 (Msolwa Station & Msolwa game) |
| **Mlimba DC** | | | | | | | |
| Mwembeni | 5 | 3332 | 43.6 | 1453 | 45 | 3 | 2 (Mlimba & private) |
| Mlimba'A' | 7 | 9485 | 43.6 | 4136 | 126 | 4 | 2 (Mlimba & private) |
| Luvirikila | 5 | 2393 | 43.6 | 1043 | 53 | 3 | 1 (Mkangawalo) |
| Mkangawalo | 3 | 5385 | 43.6 | 2348 | 118 | 2 | 1 (Mkangawalo) |
| **Mbeya region** | | | | | | | |
| **Mbeya DC** | | | | | | | |
| Igalukwa | 7 | 2670 | 45.1 | 1204 | 76 | 4 | 1 (Iwowo) |
| Iwowo | 10 | 3357 | 45.1 | 1514 | 95 | 5 | 1 (Iwowo) |
| Ikukwa | 14 | 5226 | 45.1 | 2357 | 109 | 7 | 1 (Ikukwa) |
| Simboya | 9 | 2930 | 45.1 | 1321 | 62 | 5 | 1 (Ikukwa) |
| **Mbeya CC[b]** | | | | | | | |
| Iduda | - | 979 | 39 | 382 | 77 | - | 1 (Iziwa) |
| Imbega | - | 1200 | 39 | 468 | 94 | - | 1 (Iziwa) |
| Mwambenja | - | 5981 | 39 | 2332 | 83 | - | 2 (Iganzo & SDA) |
| Nkuyu | - | 6395 | 39 | 2494 | 88 | - | 2 (Iganzo & SDA) |

[a] Population data for 2020 was estimated using a population growth rate per year, based on the population count for the base year of 2012 obtained from the national bureau of statistics.
[b] Urban councils (i.e., Mbeya CC) have only streets without further sub-divisions.

**Table S3.** Characteristics of antibiotics available in Tanzania. Antibiotics quantified are highlighted.

| Antibiotic name | Class | Spectrum | WHO category (10) | Essential medicines list (11) | Available at primary care level (11) | Available at pharmacies (12) | Half-life (hrs) |
|---|---|---|---|---|---|---|---|
| Amikacin(13) | Aminoglycosides | Broad | Access | Yes | No | Yes | 2.2 – 2.4 |
| Amoxicillin (14) | Penicillins | Broad | Access | Yes | Yes | Yes | 1 |
| Ampicillin (15) | Penicillins | Broad | Access | Yes | Yes | Yes | 0.77 |
| Ampicillin + Sulbactum (15) | Penicillins/BLIs | Broad | Access | Yes | No | Yes | 0.8 |
| Azithromycin (16) | Macrolides | Broad | Watch | Yes | Yes | Yes | 11 - 14 |
| Benzathine benzyl penicillin | Penicillins | Narrow | Access | Yes | Yes | Yes | 336 |
| Penicillin G | Penicillins | Narrow | Access | Yes | Yes | Yes | 0.30 |
| Cefaclor (17) | Cephalosporins | Broad | Watch | No | No | Yes | 1 – 2 |
| Cefadroxil (17) | Cephalosporins | Broad | Access | No | No | Yes | 1 – 2 |
| Cefazolin (17) | Cephalosporins | Broad | Access | No | No | Yes | 1 – 2 |
| Cefepime (17) | Cephalosporins | Broad | Watch | Yes | No | Yes | 1 – 2 |
| Cefixime (17) | Cephalosporins | Broad | Watch | No | No | Yes | 1 – 2 |
| Cefoperazone + Salbactum (17) | Cephalosporins/BLIs | Broad | Watch | No | No | Yes | 1 – 2 |
| Cefotaxime (17) | Cephalosporins | Broad | Watch | No | No | Yes | 1 – 2 |
| Cefotetan (17) | Cephalosporins | Broad | Watch | No | No | Yes | 3.5 |
| Cefoxitin (17) | Cephalosporins | Broad | Watch | No | No | Yes | 1 – 2 |
| Cefpirome (17) | Cephalosporins | Broad | Watch | No | No | Yes | 1 – 2 |
| Cefprozil (17) | Cephalosporins | Broad | Watch | No | No | Yes | 1 – 2 |
| Ceftazidime (18) | Cephalosporins | Broad | Watch | Yes | No | Yes | 1-3 |
| Ceftriaxone(18) | Cephalosporins | Broad | Watch | Yes | Yes | Yes | 8 – 10 |
| Ceftriaxone+sulbactam(17) | Cephalosporins | Broad | Watch | Yes | No | Yes | 8 – 10 |
| Cefuroxime (17) | Cephalosporins | Broad | Watch | Yes | No | Yes | 3-5 |
| Cephalexin (17) | Cephalosporins | Broad | Access | Yes | Yes | Yes | 1 – 2 |
| Chloramphenicol (19) | Phenicol | Broad | Access | Yes | Yes | Yes | 1- 12 |
| Ciprofloxacin (20) | Fluoroquinolone | Broad | Watch | Yes | Yes | Yes | 3-5 |
| Clarithromycin (16) | Macrolides | Broad | Watch | Yes | No | Yes | 2.6 – 2.7 |
| Amoxiclav (14) | beta-lactam | Broad | Access | Yes | Yes | Yes | 1 |
| Clindamycin (21) | lincomycin | Broad | Access | Yes | No | Yes | 2 – 2.5 |
| Cloxacillin (22) | Penicillins | Broad | Access | Yes | Yes | Yes | 1 |
| Colistin (23) | Polypeptides | Broad | Reserve | Yes | No | Yes | 1-4 |
| Dapsone (24) | sulfonyldianiline | Broad | Watch | Yes | No | Yes | 1.1 |
| Doripenem (25) | Carbapenems | Broad | Watch | No | No | Yes | 1 |
| Doxycycline (26) | Tetracyclines | Broad | Access | Yes | Yes | Yes | 18-22 |
| Ertapenem (27) | Carbapenems | Broad | Watch | No | No | Yes | 3.8 - 4.4 |
| Erythromycin (16) | Macrolides | Broad | Watch | Yes | Yes | Yes | 3.9 – 4.2 |
| Flucloxacillin (28) | Penicillins | Narrow | Access | Yes | No | Yes | 1.31-1.39 |
| Gentamicin(13) | Aminoglycosides | Broad | Access | Yes | Yes | Yes | 1.7 – 4.9 |
| Imipenem/cilastatin (29) | Carbapenems | Broad | Watch | No | No | Yes | 1 |
| Kanamycin(13) | Aminoglycosides | Broad | Watch | Yes | No | Yes | 2.1 – 2.4 |
| Levofloxacin (30) | Fluoroquinolone | Broad | Watch | Yes | No | Yes | 6-8 |
| Meropenem (31) | Carbapenems | Broad | Watch | Yes | No | Yes | 1 |
| Metronidazole (32) | Imidazole | Broad | Access | Yes | No | Yes | 8.8-10.5 |
| Moxifloxacin (33) | Fluoroquinolone | Broad | Watch | Yes | No | Yes | 8-15 |
| Neomycin | Aminoglycosides | Broad | Watch | No | No | Yes | - |
| Nitrofurantoin (34) | Macrodantin | Broad | Access | Yes | Yes | Yes | 0.3–1 |
| Norfloxacin (20) | Fluoroquinolone | Broad | Watch | No | No | Yes | 3 – 5 |
| Ofloxacin (35) | Fluoroquinolone | Broad | Watch | No | No | Yes | 5 – 8 |
| Oxytetracycline | Tetracyclines | Broad | Watch | Yes | Yes | Yes | - |
| Penicillin V (36) | Penicillins | Broad | Access | Yes | Yes | Yes | 1.5 – 2 |
| Piperacillin + tazobactam (36) | Penicillins/BLIs | Broad | Watch | Yes | No | Yes | 0.7-1 |
| Plazomicin | Aminoglycosides | Broad | Reserve | No | No | Yes | 3.5 |
| Streptomycin (37) | Aminoglycosides | Broad | Watch | No | No | Yes | 2.5 |
| Sulfamethoxazole + trimethoprim (38) | Trimethoprim /Sulfonamides | Broad | Watch | Yes | Yes | Yes | 9-15 |
| Sulfasalazine(38) | Sulfonamides | Broad | Access | No | No | Yes | 9-15 |
| Tetracycline (39) | Tetracyclines | Broad | Access | Yes | No | Yes | 8-25 |
| Tobramycin (40) | Aminoglycosides | Broad | Watch | No | No | Yes | 3 |
| Vancomycin (41) | Glycopeptides & lipoglycopeptide | Broad | Watch | Yes | No | Yes | 3 – 9 |

**Table S4.** Child level variables that were measured in the survey.

| Variable | Type | Definition | Prevalence of antibiotics in blood | Used in regression | Justification |
|---|---|---|---|---|---|
| **Sex** | Binary | Child's sex | | Yes | Typical covariate to include |
| Male | | | 147/865 (17.0) | | |
| Female | | | 149/834 (17.9) | | |
| **Age** | Categorical | Child's age in years, categorized | | Yes | Typical covariate to include; related to antibiotic prevalence |
| < 5 | | | 159/724 (22.0) | | |
| 5-9 | | | 85/598 (14.2) | | |
| 10 to 14 | | | 52/377 (13.8) | | |
| **Relationship of HoHH** | Categorical | Relationship of the head of household to the surveyed child | | No | Not related to antibiotic prevalence |
| Biological parent | | | 190/1123 (16.9) | | |
| Other | | | 106/576 (18.4) | | |
| **Exempted from paying for healthcare** | Binary | If child receives healthcare services free of charge (no, yes) | | Yes | Related to antibiotic prevalence |
| No | | | 175/1156 (15.1) | | |
| Yes | | | 121/543 (22.3) | | |
| **Valid health insurance** | Binary | Any nationally recognized, not expired health insurance card | | No | Not related to antibiotic prevalence and rare |
| No | | | 285/1623 (17.6) | | |
| Yes | | | 11/76 (14.5) | | |
| **Illness in the last 14 days** | Binary | Self-reported if child had any illness within 14 days of the survey | | Yes | Related to antibiotic prevalence |
| No | | | 182/1187 (15.3) | | |
| Yes | | | 114/512 (22.3) | | |
| **Illness type** | Categorical | Type of illness reported, composed of multiple variables (others include headache, urinary, skin, ear, nose, mouth and throat, anemia, gout) | | No<br>Sub-analysis | Small sample |
| Fever only | | | 109/512 (21.2) | | |
| GI symptoms only | | | 49/512 (9.57) | | |
| Respiratory symptoms only | | | 207/512 (40.4) | | |
| Other symptoms only | | | 29/512 (5.66) | | |
| Fever + another symptom | | | 64/512 (12.5) | | |
| Other combination of symptoms | | | 54/512 (10.5) | | |
| **Sought care for illness** | Binary | If child sought care from any place within 14 days of the survey | | No<br>Sub-analysis | Small sample |
| No | | | 200/512 (39.1) | | |
| Yes | | | 312/512 (60.9) | | |
| **Place of care seeking** | Categorical | Place where the sick child sought care (others include friend or neighbor, CHW, traditional healer, etc.) | | No<br>Sub-analysis | Small sample |
| Health facility only | | | 181/312 (58.0) | | |
| Pharmacy/drug dispensary only | | | 105/312 (33.6) | | |
| Health facility and pharmacy | | | 15/312 (4.80) | | |
| Other place | | | 11/312 (3.52) | | |
| **Malaria test result** | Categorical | Result of the malaria test conducted when seeking care | | No<br>Sub-analysis | Small sample |
| Positive | | | 43/312 (13.7) | | |
| Negative | | | 49/312 (15.7) | | |
| Don't remember | | | 4/312 (1.28) | | |
| Not performed | | | 216/312 (69.2) | | |
| **Antibiotics received** | Binary | If the child received any antibiotic when seeking care or additionally | | No<br>Sub-analysis | Small sample |
| No | | | 180/312 (57.7) | | |
| Yes | | | 132/312 (42.3) | | |

| Variable | Type | Definition | Prevalence of antibiotics in blood | Used in regression | Justification |
|---|---|---|---|---|---|
| **Type of antibiotic received** | Binary | If the child | | No | Small sample |
| Amoxicillin | | received any of the | 45/132 (34.1) | Sub-analysis | |
| Metronidazole | | specific antibiotics | 24/132 (18.2) | | |
| Ceftriaxone | | when seeking care | 0/132 (0) | | |
| Cotrimoxazole | | or additionally | 28/132 (21.2) | | |
| Cephalexin | | | 1/132 (0.75) | | |
| Ciproflaxacin | | | 6/132 (4.54) | | |
| Doxycycline | | | 0/132 (0) | | |
| Chloramphenicol | | | 2/132 (1.51) | | |
| Erythromycin | | | 6/132 (4.54) | | |
| Penicillin V | | | 5/132 (3.78) | | |
| Penicillin G | | | 0/132 (0) | | |
| Amoxicillin/clavulanic acid | | | 0/132 (0) | | |
| Azithromycin | | | 1/132 (0.75) | | |
| Ampicillin | | | 13/132 (9.84) | | |
| Gentamicin | | | 3/132 (2.27) | | |
| Ampicloxacillin | | | 7/132 (5.30) | | |
| **Taken antibiotics in the last 14 days** | Binary | Self-reported use antibiotics from | | Yes | Related to antibiotic prevalence |
| No | | any source within | 238/1548 (15.4) | | |
| Yes | | 14 days of the survey | 58/151 (38.4) | | |
| **Type of antibiotic taken** | Binary | If the child's | | No | Small sample |
| Amoxicillin | | caregiver self- | 54/151 (35.7) | Sub-analysis | |
| Metronidazole | | reported giving | 28/151 (18.5) | | |
| Ceftriaxone | | any of the specific | 0/151 (0) | | |
| Cotrimoxazole | | antibiotics | 31/151 (20.5) | | |
| Cephalexin | | (regardless of | 1/151 (0.66) | | |
| Ciproflaxacin | | illness and care | 6/151 (3.97) | | |
| Doxycycline | | seeking status) | 0/151 (0) | | |
| Chloramphenicol | | | 2/151 (1.32) | | |
| Erythromycin | | | 9/151 (5.96) | | |
| Penicillin V | | | 6/151 (3.97) | | |
| Penicillin G | | | 0/151 (0) | | |
| Amoxicillin/clavulanic acid | | | 0/151 (0) | | |
| Azithromycin | | | 3/151 (1.98) | | |
| Ampicillin | | | 13/151 (8.60) | | |
| Gentamicin | | | 3/151 (1.98) | | |
| Ampicloxacillin | | | 7/151 (4.63) | | |
| **mRDT result** | Categorical | Rapid malaria | | Yes | Related to antibiotic prevalence |
| Negative | | diagnostic test | 255/ 1425 (17.9) | | |
| Positive | | results | 41 / 274 (15.0) | | |
| **Antibiotic present in DBS sample** | Binary | If any antibiotic was detected in the | | Yes | Main outcome variable |
| No | | child's blood | 1448/1699 (85.2) | | |
| Yes | | sample | 251/1699 (14.8) | | |
| **Type of antibiotic present** | Categorical | | | No | Small sample |
| Amoxicillin | | | 43 (2.5) | Sub-analysis | |
| Metronidazole | | | 61 (3.6) | | |
| Ceftriaxone | | | 1 (0.06) | | |
| Cephalexin | | | 10 (0.6) | | |
| Ciproflaxacin | | | 16 (0.9) | | |
| Doxycycline | | | 1 (0.06) | | |
| Chloramphenicol | | | 2 (0.1) | | |
| Erythromycin | | | 29 (1.7) | | |
| Penicillin V | | | 1 (0.06) | | |
| Penicillin G | | | 1 (0.06) | | |
| Azithromycin | | | 20 (1.2) | | |
| Ampicillin | | | 13 (0.8) | | |
| Cloxacillin | | | 12 (0.7) | | |
| Sulfamethoxazol | | | 102 (6.0) | | |
| Trimethoprim | | | 144 (8.4) | | |

**Table S5.** Household-level variables that were measured in the survey (excluding SES variables).

| Variable | Type | Definition | Prevalence of antibiotics in blood | Used in regression | Justification |
|---|---|---|---|---|---|
| **No. of HH members[a]** | Categorical | Total number of people living in the household | | No | Represented in SES variable |
| <5 | | | 72/391 (18.4) | | |
| 5-7 | | | 137/881 (15.6) | | |
| 8+ | | | 87/427 (20.4) | | |
| **No. of children <15** | Categorical | Number of children under 15 years in the household | | No | Represented in SES variable |
| 1-2 | | | 103/651 (15.8) | | |
| 3-4 | | | 114/658 (17.3) | | |
| 5+ | | | 79/390 (20.3) | | |
| **No. of bedrooms[a]** | Categorical | Total number of bedrooms in the household | | No | Represented in SES variable |
| 1-2 | | | 140/837 (16.7) | | |
| 3-4 | | | 130/709 (18.3) | | |
| 5+ | | | 26/153 (17.0) | | |
| **Crowding** | Categorical | Calculated: number of people per room | | No | Represented in SES variable |
| Not crowded (≤2) | | | 108/645 (16.7) | | |
| Crowded (3-5) | | | 104/585 (17.8) | | |
| Overcrowded (6+) | | | 84/469 (17.9) | | |
| **HoHH sex** | Categorical | Sex of the head of household | | No | Not related to antibiotic prevalence |
| Male | | | 220/1222 (18.0) | | |
| Female | | | 76/477 (15.9) | | |
| **HoHH age** | Categorical | Age of the head of household | | No | Not related to antibiotic prevalence |
| <35 | | | 68/379 (17.9) | | |
| 35-49 | | | 143/787 (18.2) | | |
| 50+ | | | 85/533 (15.9) | | |
| **HoHH marital status** | Categorical | Marital status of head of household | | No | Not related to antibiotic prevalence |
| Married | | | 143/868 (16.5) | | |
| Living together | | | 95/547 (17.4) | | |
| Divorced/separated/widowed/ Single | | | 58/284 (20.4) | | |
| **HoHH education level** | Categorical | Highest level of education attained by the head of household | | No | Represented in SES variable |
| No education | | | 42/312 (13.5) | | |
| Primary | | | 187/1240 (15.1) | | |
| Vocational/O-level/higher | | | 22/143 (15.4) | | |
| **HoHH employment** | Categorical | Main economic activity of the head of household. | | No | Represented in SES variable |
| Any other type of employment | | | 48/245 (19.5) | | |
| Agriculture | | | 196/1412 (13.8) | | |
| Unemployed (retired, housewife, unable to work, etc.) | | | 7/39 (17.9) | | |
| **SES quintile[b]** | Categorical | Socioeconomic status quintile | | Yes | Related to antibiotic prevalence |
| 1 | | | 47/340 (13.8) | | |
| 2 | | | 60/402 (14.9) | | |
| 3 | | | 53/277 (19.1) | | |
| 4 | | | 68/341 (19.9) | | |
| 5 | | | 68/339 (20.1)) | | |
| **Usual place of care** | Categorical | Usual place children are taken when sick | | Yes | Related to antibiotic prevalence |
| Health facility | | | 235/1586 (14.8) | | |
| Drug shop or other | | | 10/113 (8.8) | | |
| **Travel time to place of care** | Categorical | Time needed to reach the usual place of care | | Yes | Related to antibiotic prevalence |
| <15 min | | | 83/501 (16.5) | | |
| 15 min – 1 hr | | | 129/859 (15.0) | | |
| >1 hr | | | 39/339 (11.5) | | |
| **Transport to place of care** | Categorical | Typical mode of transport taken to reach the usual place of care. | | No | Not related to antibiotic prevalence |
| Foot | | | 238/1332 (17.9) | | |
| Other (bicycle, motorcycle, public transport or car) | | | 58/367 (15.8) | | |
| **Store medicines at home** | Binary | Reported storage of any medicines at home | | Yes | Related to antibiotic prevalence |
| No | | | 193/1299 (14.9) | | |
| Yes | | | 103/400 (25.8) | | |
| **Store antibiotics at home** | Binary | If the ATB were found in the HH | | Yes | Related to ATB prevalence |
| No | | | 223/1503 (14.8) | | |
| Yes | | | 73/196 (37.2) | | |

| Variable | Type | Definition | Prevalence of antibiotics in blood | Used in regression | Justification |
|---|---|---|---|---|---|
| **Store specific antibiotics** | | | | Sub-analysis | Small sample |
| Amoxicillin | | | 55 | | |
| Metronidazole | | | 43 | | |
| Ceftriaxone | | | 0 | | |
| Cotrimoxazole | | | 42 | | |
| Cephalexin | | | 8 | | |
| Ciproflaxacin | | | 6 | | |
| Doxycycline | | | 2 | | |
| Chloramphenicol | | | 7 | | |
| Erythromicin | | | 27 | | |
| Penicillin V | | | 4 | | |
| Penicillin G | | | 0 | | |
| Amoxicillin/clavulanic acid | | | 8 | | |
| Azithromycin | | | 15 | | |
| Ampicillin | | | 27 | | |
| Gentamicin | | | 0 | | |
| Ampicloxacillin | | | 7 | | |
| **Used stored medicines** | Binary | If caregiver has ever given stored medicine to child <15 | | No | Not related to antibiotic prevalence |
| No | | | 172/1223 (14) | | |
| Yes | | | 79/476 (16.6) | | |

[a] These two variables were used to calculate crowding status as number of household members per bedroom

[b] SES quintile was determined based on the variables and method summarized in Table S5

**Table S6:** Socioeconomic variables

| Variable | Order (value) | Distribution |
|---|---|---|
| **Crowding status**[a] | | |
| Overcrowded (6+) | 1 | 469 |
| Crowded (3-5) | 2 | 585 |
| Not crowded (≤2) | 3 | 645 |
| **HoHH education level**** | | |
| No education | 1 | 312 |
| Primary | 2 | 1240 |
| Vocational | 3 | 10 |
| Secondary | 4 | 117 |
| College or university | 5 | 16 |
| **HoHH employment** | | |
| Unemployed (including retired, housewife) | 1 | 58 |
| Agriculture/farming | 2 | 1412 |
| Animal breeder/fisherman | 3 | 19 |
| Small business/street trader | 4 | 80 |
| Business (driver, livestock, charcoal, etc.) | 5 | 51 |
| Domestic or casual worker | 6 | 24 |
| Employee or skilled worker | 7 | 55 |
| **Floor type**** | | |
| Dung | 1 | 4 |
| Earth | 2 | 995 |
| Wood | 3 | 670 |
| Cement | 4 | 25 |
| Ceramic | 5 | 5 |
| **Wall type**** | | |
| Grass, boxes | 1 | 47 |
| Tin, clay, wood | 2 | 152 |
| Raw bricks (w or w/o plaster) | 3 | 325 |
| Stones, cement, burnt bricks | 4 | 1175 |
| **Cooking fuel type** | | |
| Wood (+straw/shrubs) | 1 | 1263 |
| Charcoal | 2 | 375 |
| Electricity/solar/gas/kerosene | 3 | 61 |
| **Tin or tiled roof**** | | |
| No | 0 | 192 |
| Yes | 1 | 1507 |
| **Presence of electricity** | | |
| No | 0 | 980 |
| Yes | 1 | 719 |
| **Toilet type**** | | |
| Unimproved | 1 | 813 |
| Improved used by multiple HH | 2 | 130 |
| Improved used by one HH | 3 | 756 |
| **Water source type** | | |
| River | 1 | 164 |
| Open well | 2 | 290 |
| Spring | 3 | 89 |
| Dug well | 4 | 246 |
| Tap | 5 | 910 |
| **Time to water source** | | |
| On premises | 4 | 40 |
| Less than 15 min | 3 | 290 |
| 15-30 min | 2 | 993 |
| More than 30 min | 1 | 376 |

| | | |
|---|---|---|
| **Transport ownership** | | |
| None | 1 | 759 |
| Bicycle only | 2 | 651 |
| Motorbike (w or w/o bicycle) | 3 | 268 |
| Car (w or w/o bicycle or motorbike) | 4 | 21 |
| **Fridge ownership** | | |
| No | 0 | 1629 |
| Yes | 1 | 70 |
| **Milling & sewing machine ownership** | | |
| None | 1 | 1556 |
| Sewing machine only | 2 | 121 |
| Milling machine (w or w/o sewing machine) | 3 | 22 |
| **Telecom ownership**** | | |
| None | 1 | 715 |
| Radio only | 2 | 607 |
| TV (w or w/o satellite dish) | 3 | 377 |
| **Iron ownership**** | | |
| No | 0 | 1298 |
| Yes | 1 | 401 |
| **Phone ownership** | | |
| No phone | 1 | 250 |
| Regular cell phone only | 2 | 1196 |
| Smart phone | 3 | 253 |
| **Watch ownership** | | |
| No | 0 | 1403 |
| Yes | 1 | 296 |
| **Livestock worth (USD)[b]** | | |
| 0 | 1 | 520 |
| 1-100 | 2 | 678 |
| 101-250 | 3 | 206 |
| 251-500 | 4 | 131 |
| 501-1000 | 5 | 53 |
| 1001-5000 | 6 | 85 |
| >5000 | 7 | 26 |

[a] calculated by dividing the number of household members by the number of bedrooms and categorized
[b] calculated by multiplying the number of each type of livestock owned by the household by the value of each animal (below).

** Variables were included in the final SES score.

| Type of animal | Value in TZS (USD) |
|---|---|
| No. of cows | 400,000 (172) |
| No. of goats | 50,000 (22) |
| No. of sheep | 60,000 (26) |
| No. of pigs | 100,000 (43) |
| No. of rabbits | 10,000 (4.5) |
| No. of guinea pigs | 6,000 (2.5) |
| No. of chickens | 15,000 (6.5) |
| No. guinea fowl | 20,000 (8.5) |
| No. of ducks | 20,000 (8.5) |
| No. of pigeons | 2,000 (1) |

**Table S7.** Summary of 27 children who had three or more antibiotics in their blood. Blue [1] indicates presence of antibiotic in blood sample. Y indicates presence of risk factor. Red indicates absence of all four risk factors.

| Region | Ward | Age | Trimethoprim | Sulfamethoxazol | Metronidazole | Amoxicillin | Erythromycin | Azithromycin | Ciprofloxacin | Ampicillin | Cloxacillin | Cephalexin | Chloramphenicol | Penicillin V | Penicillin G | Doxycycline | Ceftriaxone | Illness in last 14 days | Sought care for illness | Reported taking antibiotic | Antibiotic found at home | Place of care | Antibiotics reported taking | Antibiotics found at home |
|---|---|---|---|---|---|---|---|---|---|---|---|---|---|---|---|---|---|---|---|---|---|---|---|---|
| mbeya | iziwa | 5-9 | | 1 | 1 | | | 1 | 1 | | 1 | 1 | | | | 1 | | | | | | | | |
| mbeya | iganzo | <5 | 1 | 1 | 1 | 1 | | 1 | | | | | | | | | | | | Y | Y | | Azithromycin | Azithromycin |
| morogoro | mlimba | 10-14 | 1 | 1 | 1 | | | | | | | 1 | | | | | | Y | Y | Y | Y | Health facility | Cotrimoxazole | Cotrimoxazole |
| morogoro | kidatu | 5-9 | 1 | 1 | | | 1 | 1 | | | | | | | | | | | | | | | | |
| morogoro | mngeta | 10-14 | 1 | 1 | | | | | 1 | | | | | | | | | | | | Y | | | Cotrimoxazole |
| morogoro | mlimba | 10-14 | 1 | 1 | 1 | | | | | | | | | | | | | | | | | | | |
| mbeya | iganzo | 10-14 | 1 | 1 | 1 | | | | | | | | | | | | | | | Y | Y | | Metronidazole | Metronidazole |
| mbeya | iganzo | 5-9 | 1 | 1 | | | | 1 | | | | | | | | | | | | | | | | |
| mbeya | iganzo | <5 | 1 | 1 | 1 | | | | | | | | | | | | | Y | Y | | Y | Health facility & drug shop | | Metronidazole |
| mbeya | iziwa | <5 | | 1 | | | | | | 1 | 1 | | | | | | | | | | | | | |
| mbeya | iziwa | <5 | 1 | | | | | | | 1 | 1 | | | | | | | Y | | | | | | |
| mbeya | iganzo | 5-9 | 1 | 1 | 1 | | | | | | | | | | | | | | | | | | | |
| morogoro | mngeta | <5 | 1 | 1 | | | | | | 1 | | | | | | | | Y | Y | Y | | Health facility | Ampicillin | |
| morogoro | mngeta | 10-14 | 1 | 1 | | | 1 | | | | | | | | | | | Y | | | | | | |
| morogoro | mngeta | 5-9 | 1 | 1 | | | | | 1 | | | | | | | | | Y | Y | | | Drug shop | | |
| mbeya | itawa | 5-9 | 1 | 1 | | | 1 | | | | | | | | | | | Y | Y | | | Health facility | | |
| mbeya | iganzo | 10-14 | 1 | 1 | 1 | | | | | | | | | | | | | | | Y | Y | | Metronidazole | Metronidazole, amoxiclav |
| morogoro | euga | <5 | 1 | 1 | 1 | | | | | | | | | | | | | Y | Y | | | Health facility | | |
| morogoro | kidatu | <5 | 1 | 1 | | 1 | | | | | | | | | | | | | | | | | | |
| mbeya | ikukwa | <5 | 1 | 1 | | | 1 | | | | | | | | | | | Y | Y | | | Health facility & drug shop | | |
| mbeya | itawa | <5 | 1 | 1 | | | 1 | | | | | | | | | | | Y | Y | Y | Y | Health facility | Cotrimoxazole | Metronidazole |
| morogoro | euga | <5 | 1 | 1 | | | | | | | | | 1 | | | | | | | Y | | | Cotrimoxazole | |
| mbeya | ikukwa | <5 | 1 | 1 | 1 | | | | | | | | | | | | | Y | Y | | | Health facility | | |
| mbeya | ikukwa | 5-9 | 1 | 1 | | 1 | | | | | | | | | | | | Y | Y | Y | | Health facility & drug shop | Gentamicin | |
| mbeya | ikukwa | <5 | 1 | 1 | | 1 | | | | | | | | | | | | Y | Y | Y | | Health facility | Cotrimoxazole | |
| morogoro | mlimba | <5 | 1 | 1 | | | | | | | | 1 | | | | | | Y | | Y | Y | | Cotrimoxazole | Cephalexin, erythromycin |
| mbeya | iziwa | <5 | 1 | 1 | | | 1 | | | | | | | | | | | Y | Y | Y | Y | Health facility | Erythromycin | Azithromycin |

**Table S8.** Summary N (%) of variables collected on the subset of children (N=512) who had experienced illness in the 14 days prior to the survey and univariable and multivariable regression model results explaining presence of antibiotics in the blood.

| | Mbeya | Morogoro | Overall | Univariable OR (95% CI) | p-value | Multivariable aOR (95% CI) | p-value |
|---|---|---|---|---|---|---|---|
| **Illness type*** | **N=235** | **N=277** | **N=512** | **N=512** | | **N=312** | |
| Fever only | 22 (9.4) | 87 (31.4) | 109 (21.3) | 1.00 | | 1.00 | |
| Gastrointestinal (GI) symptoms only | 31 (13.2) | 18 (6.50) | 49 (9.60) | 1.38 (0.60–3.17) | 0.448 | 2.11 (0.74–6.05) | 0.165 |
| Respiratory symptoms only | 123 (52.3) | 84 (30.3) | 207 (40.4) | 1.16 (0.63–2.15) | 0.634 | 1.50 (0.68–3.32) | 0.316 |
| Other symptom only[a] | 11 (4.70) | 18 (6.50) | 29 (5.70) | 0.92 (0.31–2.74) | 0.881 | 1.35 (0.38–4.82) | 0.643 |
| Fever + GI or respiratory (regardless of other symptoms) | 28 (11.9) | 36 (13.0) | 64 (12.5) | 1.89 (0.90–3.99) | 0.093 | 1.48 (0.60–3.65) | 0.395 |
| Any other combination of multiple symptoms | 20 (8.50) | 34 (12.3) | 54 (10.5) | 1.28 (0.57–2.85) | 0.546 | 1.01 (0.38–2.68) | 0.992 |
| **Sought care#** | **N=235** | **N=277** | **N=512** | **N=512** | | | |
| No | 102 (43.4) | 98 (35.4) | 200 (39.1) | 1.00 | | **---** | --- |
| Yes | 133 (56.6) | 179 (64.6) | 312 (60.9) | 2.36 (1.46–3.84) | **0.001** | | |
| **Place sought care** | **N=133** | **N=179** | **N=312** | **N=312** | | **N=312** | |
| Health facility | 88 (66.1) | 108 (60.3) | 196 (62.8) | 1.00 | | 1.00 | |
| Pharmacy, drug shop or other[b] | 45 (33.9) | 71 (39.7) | 116 (37.2) | 0.72 (0.42–1.25) | 0.240 | 0.90 (0.46–1.75) | 0.757 |
| **Malaria RDT result*** | **N=133** | **N=179** | **N=312** | **N=312** | | **N=312** | |
| Negative | 16 (12.0) | 33 (18.4) | 49 (15.7) | 1.00 | | 1.00 | |
| Positive | 2 (1.50) | 41 (22.9) | 43 (13.8) | 0.75 (0.27–2.12) | 0.593 | 1.16 (0.39–3.47) | 0.795 |
| Not performed (or don't remember) | 115 (86.5) | 105 (58.7) | 220 (70.5) | 0.59 (0.29–1.17) | 0.129 | 0.64 (0.29–1.43) | 0.281 |
| **Antibiotics received*** | **N=133** | **N=179** | **N=312** | **N=312** | | **N=312** | |
| No | 65 (48.9) | 115 (64.2) | 180 (57.7) | 1.00 | | 1.00 | |
| Yes | 68 (51.1) | 64 (35.8) | 132 (42.3) | 2.24 (1.32–3.78) | **0.003** | 2.02 (1.15–3.54) | **0.015** |

[a] Other symptoms include headache, urinary, skin, ear, nose, mouth and throat, anemia, gout.
[b] Other places include neighbor or friend, community health worker, traditional healer.
* Denotes statistically significant ($p<0.05$) difference in the variable distribution between regions according to a Fisher's exact test.
### This variable was dropped from the multivariable model because all children in this subset of 312 sought care.

**Figure S1.** Map of the study area showing wards that were selected versus those that were not selected for the study. Median number of health facilities per ward is 1.5 for selected and 3 for non-selected wards. However, non-selected wards are generally larger than selected wards. In summary, selected wards represent average access to health facilities in the study area, considering their urban, peri-urban or rural status.

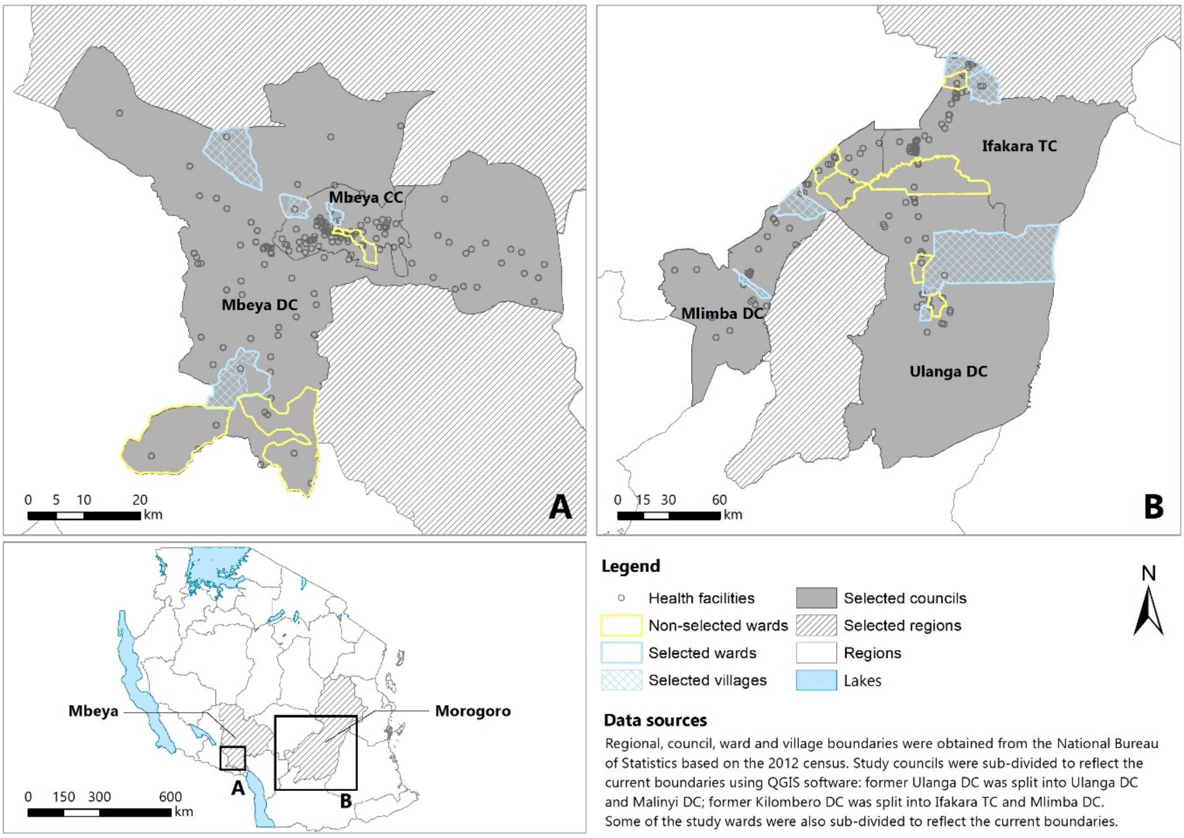
